# Systematic review of the association between rotavirus infection, or rotavirus vaccination and coeliac disease

**DOI:** 10.1101/2020.12.01.20241869

**Authors:** Thomas Inns, Kate M Fleming, Miren Iturriza-Gomara, Daniel Hungerford

## Abstract

**Background:** There is some evidence that rotavirus infection leads to an increased risk of coeliac disease (CD), and some immunological and biological plausibility for the human immune system recognising rotavirus particles and gluten proteins in a similar way. It is therefore plausible that rotavirus vaccine could have a role in preventing CD. However, such evidence has not previously been summarised in a systematic way to present a coherent picture. We conducted this systematic literature review to address this gap in the evidence. The aim of this research was to determine the nature of any association between rotavirus infection, or rotavirus vaccination, and risk of CD.

**Methods:** We searched Scopus, MEDLINE, Europe PMC and medRxiv for studies published between 01 January 1980 and 31 July 2020, using terms related to CD and rotavirus. Publications were screened independently by two reviewers using exclusion criteria. We extracted data from included papers using a standardized data extraction form and assessed risk of bias using the Newcastle-Ottawa Scale. Outcomes were descriptions of the settings and methods reported in included papers, and any estimates of effect.

**Results:** After exclusions, we reviewed five papers of which two used the exposure of rotavirus infection and three used the exposure of rotavirus vaccination. One paper found that rotavirus infection increased the risk of CD and that this was statistically significant. None of the three publications studying the association between rotavirus vaccination and CD were graded as high quality. All found a protective effect of RotaTeq^®^ rotavirus vaccination, but this was only statistically significant in two studies.

**Conclusions:** Few studies have been published on this research question. Those that have been published are not of sufficient quality and did not use comparable methods. Due to differences in study results there remains uncertainty regarding the relationship between rotavirus infection, vaccination and CD.

## Background

Coeliac Disease (CD), spelt Celiac in US English, is a chronic inflammatory intestinal disease that is induced by exposure to gluten.(1) CD manifests in most patients as a gastrointestinal illness, with typical symptoms including chronic diarrhoea, failure to thrive and abdominal distension.(2) The global prevalence of CD is estimated to be 1.4% based on serological markers,(3) with evidence from stored serum samples that CD prevalence has increased substantially over the last 50 years, but the reason for this increase is unclear.(4) In England, the incidence of diagnosed CD showed a four-fold increase, from 5.2 to 19.1 cases per 100,000 person-years, over the period 1990-2011.(5)

Rotavirus infection typically causes diarrhoea, vomiting and fever, with symptoms commonly leading to dehydration in children.(6) Prior to the introduction of rotavirus vaccination, rotavirus caused substantial global morbidity and mortality in children under five; estimated to be 453,000 deaths per year, with over 90% in developing countries.(7) Two highly effective rotavirus vaccines were licensed for use in 2006; Rotarix^®^ and RotaTeq^®^.(8) Rotarix^®^ was added to the routine immunisation program in the United Kingdom (UK) on 01 July 2013, to be given to infants in two doses, at two and three months old.(9) This has had a positive effect infectious intestinal disease in England, averting approximately 90,000 healthcare visits for acute gastroenteritis by children per year and saving an estimated £12.5 million annually in healthcare costs.(10)

There is some evidence that CD can be triggered by rotavirus infection. A study in 2006 found that multiple rotavirus infections significantly increased the risk of CD, adjusted for other risk factors.(11) This was supported by an immunological study which showed the similarity between a coeliac peptide and a key rotavirus protein.(12) Following this, one study showed that CD prevalence was significantly lower in rotavirus vaccine recipients than in those who did not receive the vaccine.(13) However, a Finnish retrospective cohort study did not find a statistically significant association between CD and rotavirus vaccination.(14) Furthermore, a recent immunology study has found that children with CD do not have significantly different immune reactivity to rotavirus proteins than children without CD, questioning the plausibility of this mechanism to trigger CD.(15)

Based on these studies, there is some evidence that rotavirus infection leads to an increased risk of CD, and that rotavirus vaccine may possibly have a role in preventing CD. However, such evidence has not previously been summarised in a systematic way to present a coherent picture. In order to address this gap in the evidence and critically review current evidence on this topic, we conducted a systematic literature review. The aim of this research was to determine the nature of any association between rotavirus infection, or rotavirus vaccination, and risk of CD.

## Methods

We used the Preferred Reporting Items for Systematic Reviews and Meta-Analyses (PRISMA) guidelines as a basis for this review.(16) The protocol for this systematic review was registered on PROSPERO (CRD42020168166).(17)

### Eligibility criteria

We included studies published between 01 January 1980 and 31 July 2020 in peer-reviewed journals or on pre-print servers. We did not apply any geographical or language restrictions. We excluded the following types of publication: reviews, letters to the editor, editorials and conference abstracts. We also excluded any studies of molecular biology.

### Information sources

We searched the following electronic databases: Scopus, MEDLINE, Europe PMC and medRxiv. The last search date was 10 September 2020.

### Search

We used the following search terms: rotavirus.ab,ti AND (coeliac or celiac).ab,ti. In Scopus, we searched within title, abstracts and keywords; in MEDLINE, we searched within titles and abstracts. We initially developed the search terms in MEDLINE and piloted them prior to selection.

### Study selection

We imported all references identified by the search strategy into the reference management programme EndNote X8.2 (Clarivate Analytics, USA). We used this software to store and de-duplicate studies. We then applied the screening criteria to the title and abstract, then the full text; this was done independently by Dr Thomas Inns and Dr Dan Hungerford. We then reconciled any differences by discussion and agreed the final studies for inclusion by consensus. We then searched the reference lists of included studies to identify relevant articles not previously included.

### Data collection process and data items

Data extraction from included papers was performed independently, in duplicate, by both reviewers. We used a standardised data extraction form to extract relevant data from included studies. We extracted the following data: year published, study design, study setting, study sample size, data collection period, follow-up duration, main exposure, main outcome, measure of effect, effect estimate, effect confidence interval.

### Summary measures

Estimates of effect were the key summary measures; these were pooled between studies where appropriate. If studies did not report an effect estimate, we calculated this from available data where possible.

### Synthesis of results

We compared the settings, methods and estimates of effect from the included studies.

### Risk of bias (individual and across studies)

We assessed included studies for risk of bias using the Newcastle-Ottawa Scale, a risk of bias assessment tool recommended by the Cochrane Collaboration.(18) This tool assesses the quality of non-randomised studies and has established face and content validity for this purpose.(19) This categorised assessment based on selection, comparability and exposure for case-control studies and outcome for cohort studies. We included findings related to study bias in the results section.

Both reviewers conducted risk of bias and quality assessment of the identified studies independently using the Newcastle-Ottawa Scale and then reconciled their findings. Discrepancies between the quality assessments of the two reviewers were discussed and agreed based on consensus.

## Results

### Study selection

Our initial searches identified 133 papers; following de-duplication this was reduced to 81 publications. After reviewing the titles and abstracts, we included 16 publications and excluded 65 publications. We reviewed the full text of these 16 publications; of these, five papers were eligible for inclusion in this systematic review (Figure). Of the 11 papers we excluded after full text review, eight were narrative reviews and three were molecular biology studies. We did not find any relevant studies on pre-print servers and no additional publications were identified through searching reference lists of included studies.

**Figure:**
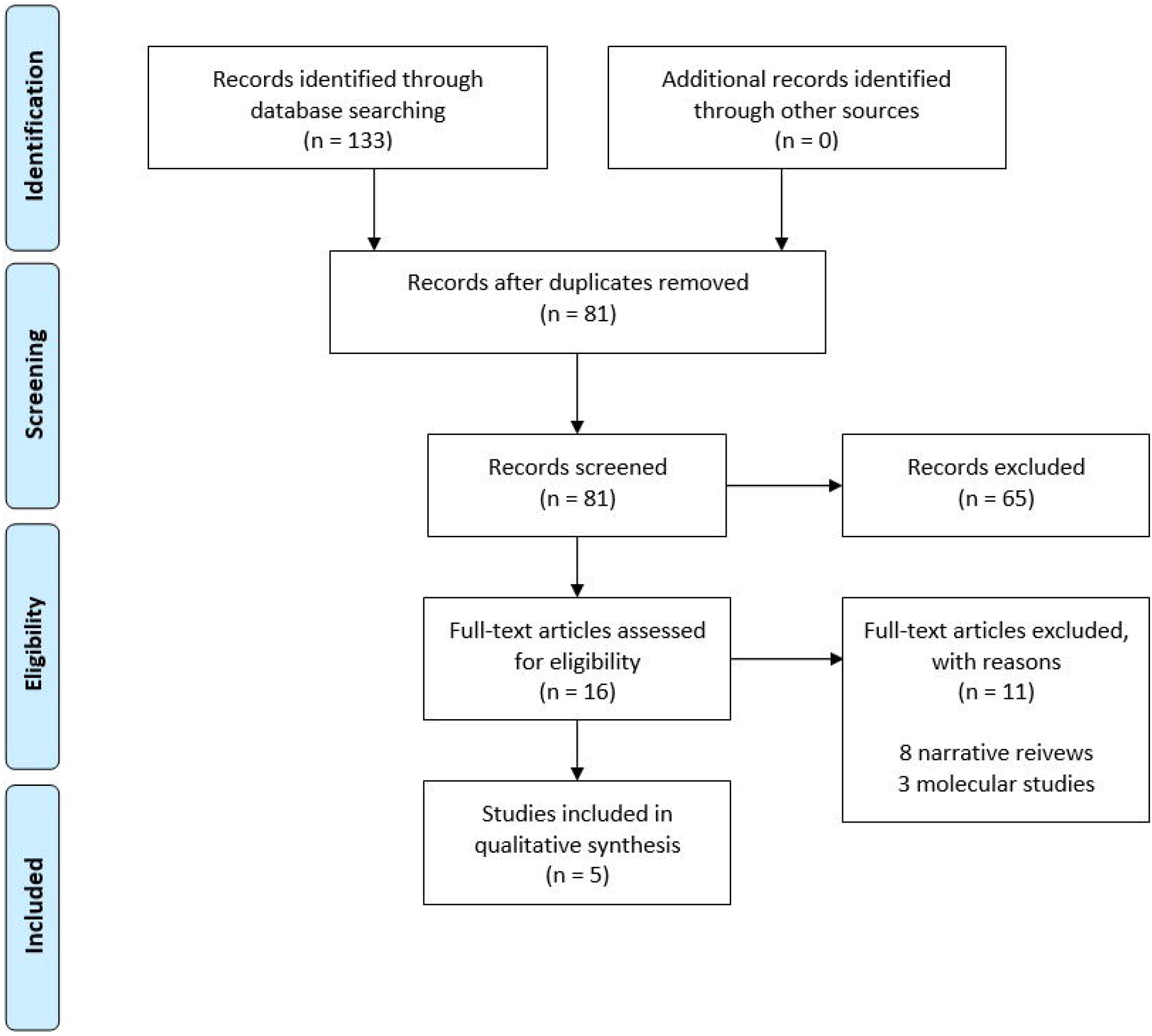
Study selection for systematic review of rotavirus infection and coeliac disease (n = 133)

### Characteristics of included studies

The first paper on this topic was published in 2006; there was a ten year gap before two papers were published in 2017, with another two papers being published in 2019. Two studies were based on populations in Finland; one study was based on a population in the state of Denver, US. The two remaining studies were based on the same sample of children in Finland, US, Germany and Sweden. For a full summary of included study characteristics, see Table 1.

**Table 1:**
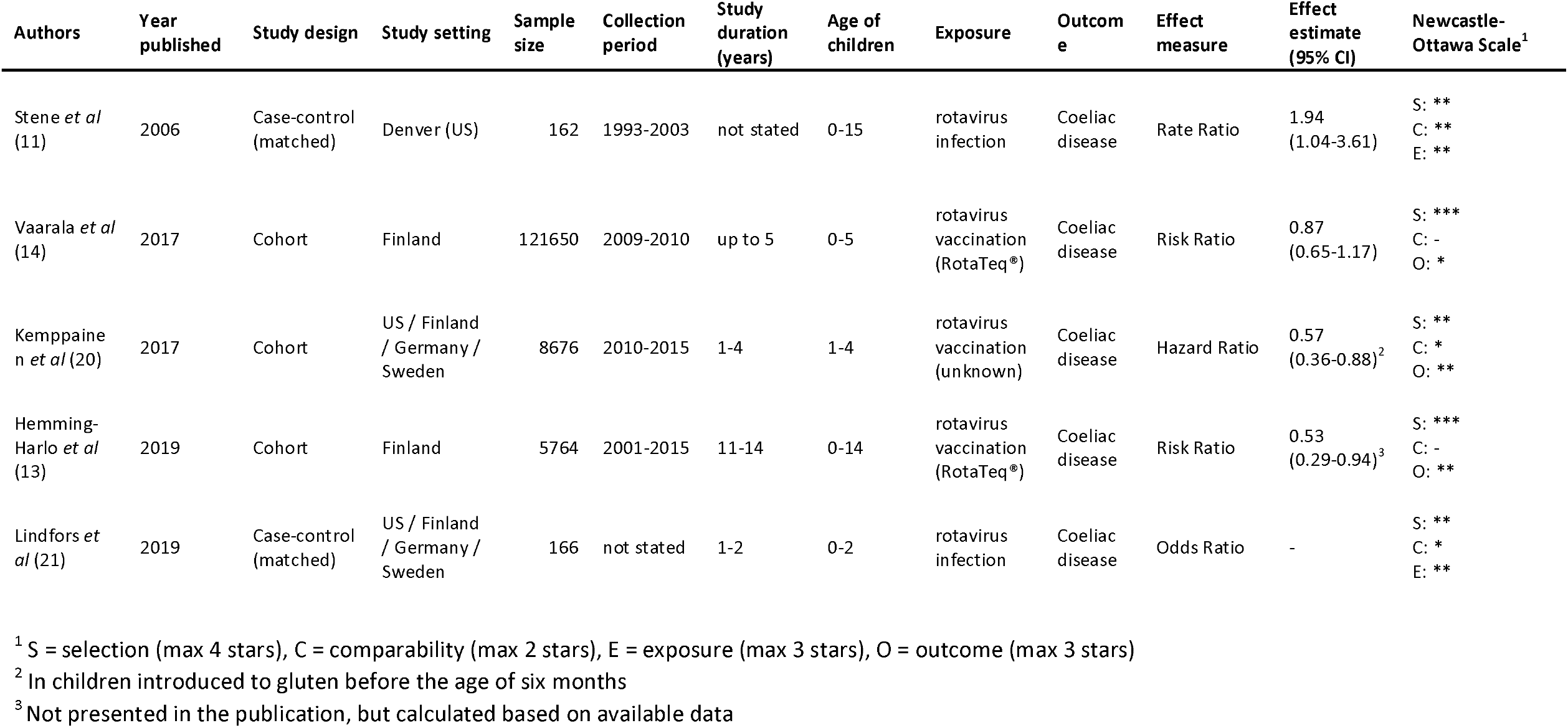
Summary of studies meeting systematic review inclusion criteria.

Three of the five publications were based on cohort study designs. The two other publications used matched case-control study designs which were nested within a defined study cohort. The two case-control studies had the smallest number of participants, with 166 and 1,931 participants. The follow-up period (one to two years) was stated only for the smaller of the two studies. Two cohort studies had larger numbers of participants, one with 5,764 and one with 8,676. The former had the longest follow-up period (11 to 14 years) and the latter had a follow-up period of one to four years. The largest cohort study had 121,650 participants with a follow-up period of up to five years.

All five studies included only children, aged between six months and 15 years old, with three of the studies being based on children known to have Human Leukocyte Antigen (HLA) alleles associated with a high risk of coeliac disease.

### Risk of bias within studies

Using the Newcastle-Ottawa scale, out of a maximum of nine, we scored the study by Stene *et al*. as six; in this study there was a lack of clarity over the control selection process. We assessed the study by Kemppainen *et al*. to be a score of five; it used unclear exposure classification and had uncertainty over elements of the statistical methods. We scored the study by Vaarala *et al*. as four; there was a lack of information on outcome ascertainment and confounder adjustment. We assessed the publications from Hemming-Harlo *et al*. and Lindfors *et al*. as five on this scale; these studies had a lack of adjustment for confounding and a lack of information on control selection, respectively.

### Results of individual studies

Two studies included in this review used rotavirus infection as the primary exposure, both were matched case-control studies. The study by Stene *et al*. estimated an increased risk of CD following rotavirus infection: a rate ratio of 1.94 (95% CI 0.39-9.56) following one rotavirus infection, and a rate ratio of 3.76 (95% CI 1.04-3.61) following two or more rotavirus infections. The study by Lindfors *et al*. was a case-control study, however no effect estimate was made as too few rotavirus infections were detected in the study sample.

Three publications in this review studied rotavirus vaccination as the primary exposure, all were cohort studies. The cohort study by Vaarala *et al*. reported a decreased risk of CD in participants who had received a rotavirus vaccination (risk ratio 0.87 [95% CI 0.65-1.17]), not statistically significant at the 5% level. This was an adjusted estimate, but as a crude estimate was not presented, it was not clear which confounders were adjusted for in the analysis. The multi-country cohort study by Kemppainen *et al*. did not report a single effect estimate for the association between rotavirus vaccination and coeliac disease or specify the exact vaccine used (depending on the country, state and year this would have either been RotaTeq^®^ or Rotarix^®^). In a subset of children introduced to gluten before six months of age, they estimated a statistically significant Hazard Ratio of 0.57 (95% CI 0.36-0.88). No effect estimate was published for children introduced to gluten after six months of age. The cohort study by Hemming-Harlo *et al*. compared differences of prevalence percentages in those vaccinated with RotaTeq^®^ and those unvaccinated. Using data presented in the paper, we calculated a decreased in risk of CD: risk ratio 0.53 (95% CI 0.29-0.94).

### Synthesis of results

Due to different study designs, different exposure variables and lack of comparable estimates of effect in the small number of individual studies, it was not appropriate to synthesise included study effect estimates in a meta-analysis.

## Discussion

### Summary of evidence

In this analysis, only one study quantified the association between rotavirus infection and increased rate of CD. This found a statistically significant association, with a rate ratio of 1.94, and evidence of increased risk from multiple infections. However, this study took place in a population of children known to have a high risk of CD, so the generalisability of these findings may be limited. Additionally, the age at which cases and controls were selected was not clear. Because older children are more likely to be diagnosed with CD,(5) this affects the confidence in inferences made around the increased risk from multiple rotavirus infections. Despite being published in 2006, no further studies have subsequently been published which replicate or refute these findings in other populations or settings. The study by Lindfors *et al* examined this question, but too few rotavirus infections were detected during the study period.(21) This lack of ascertainment of common childhood infections such as norovirus and rotavirus raises questions around the representativeness of the children involved and the stool sampling methods, given they were of an age when some these infections would be expected.

In this review, we found three studies which examined the association between rotavirus vaccination and CD. All three studies were published between 2017 and 2019, with no earlier studies being available. Two were based in Finland and one was a multi-country cohort of children known to have a high risk of CD from Finland, US, Germany and Sweden. Two studied the effect of RotaTeq^®^ vaccine and one was not clear on the vaccine used; no published studies have explicitly examined the possible association with Rotarix^®^ vaccine (or Rotavac^®^, another rotavirus vaccine). The three studies all found a protective effect of rotavirus vaccine on CD, however this relationship was only statistically significant in two studies, one of which was only in children who were exposed to gluten before six months of age. In the other study with a statistically significant result, this was a univariable association unadjusted for any confounding factors.

However, as none of these three publications scored more than five on the Newcastle-Ottawa scale, there is a substantial risk of bias in the effect estimates they present. The publication by Vaarala *et al*. does not state which confounders were adjusted for.(14) Without this information, it is possible that the presented effect was biased in either direction by not adjusting for an important confounder. The publication by Kemppainen *et al*. only presented an effect estimate for the stratum of children introduced to gluten before six months of age.(20) The lack of presented effect estimate for all children, or for those introduced to gluten after six months of age makes it difficult to interpret and raises the prospect that this may have been the result of a data-dependent analysis. In the publication by Hemming-Harlo *et al*., the authors do not present an estimate of effect or any information on confounders which would allow an adjusted estimate to be made.(13) There is also a potential selection bias in this study, arising from the differential inclusion of vaccinated children and those with autoimmune conditions, potentially biasing the association towards the null. Given that factors such as age, sex and socioeconomic status are known to be associated with both rotavirus vaccination and CD, there is a need for an effect estimate for this association which is multiply adjusted for these potential confounders.

### Limitations

This review included a small number of published studies which met the inclusion criteria. These studies were not of high quality, with none scoring more than six on the Newcastle-Ottawa scale. Common limitations among the studies included: unclear control selection, vague exposure classification, lack of outcome ascertainment information and uncertainty of statistical methods, particularly adjustment for confounders. The interpretation of these crude estimates of effect, or adjusted ones without clarity on which confounders were adjusted for, should be done with caution as confounding may affect the magnitude or direction of observed effect.

A wider limitation of the studies included in this review is that they all either include children from Finland (who are genetically at increased risk of CD), or children in developed countries known to be genetically at high risk of CD. This limits the generalisability of these findings to children in other settings, particularly those in developing countries and those without genetic risk factors for CD. It may be these other populations have a different immune response to rotavirus and therefore the protective effect of rotavirus vaccine is reduced towards null effect.

One limitation of the review design was that it was not possible to include grey literature in our search strategy due to resource limitations. This may have excluded literature such as PhD theses, official publications and conference proceedings, affecting the representativeness of our findings. Another limitation was that it was not appropriate to conduct a meta-analysis with the available results. This would have been a useful method of summarising the relationships between rotavirus infection and CD, and rotavirus vaccination and CD, had this been possible.

## Conclusions

In this systematic review, we found one published study of the association between rotavirus infection and CD; this found a statistically significant effect in a group of genetically high-risk children. There were only three studies of the association between rotavirus vaccination and CD; none were graded as high quality. All found a protective effect of rotavirus vaccination, but this was only statistically significant in two studies. Due to different study methods it was not possible to combine these estimates in a meta-analysis. There remains uncertainty regarding the relationship between infection and rotavirus vaccination and CD.

## Data Availability

All data generated or analysed during this study are included in this published article.

## List of abbreviations

CD: Coeliac Disease
CI: Confidence Interval
HLA: Human Leukocyte Antigen
PRISMA: Preferred Reporting Items for Systematic Reviews and Meta-Analyses
UK: United Kingdom

## Declarations

### Ethics approval and consent to participate

Not applicable.

### Consent for publication

Not applicable.

### Availability of data and materials

All data generated or analysed during this study are included in this published article.

### Competing interests

DH has received research grant support on the topic of rotavirus vaccines from GlaxoSmithKline Biologicals, Sanofi Pasteur and Merck and Co (Kenilworth, New Jersey, US) after the closure of Sanofi Pasteur-MSD in December 2016. The other authors declare that they have no competing interests.

### Funding

Daniel Hungerford is funded by a National Institute for Health Research (NIHR) Post-doctoral Fellowship for this research project. Daniel Hungerford and Thomas Inns are affiliated to the National Institute for Health Research Health Protection Research Unit (NIHR HPRU) in Gastrointestinal Infections at University of Liverpool in partnership with Public Health England (PHE), in collaboration with University of Warwick. Daniel Hungerford and Thomas Inns are based at the University of Liverpool. The views expressed are those of the author(s) and not necessarily those of the NHS, the NIHR, the Department of Health and Social Care or Public Health England.

### Author’s contributions

TI, KMF, MIG and DH all contributed to the study design. TI and DH undertook the search, selection and analysis. TI wrote the first manuscript draft. All authors read, contributed to and approved the final manuscript.

## Acknowledgements

Not applicable.

